# Probable transmission of SARS-CoV-2 from an African lion to zoo employees

**DOI:** 10.1101/2023.01.29.23285159

**Authors:** Audrey A. Siegrist, Kira L. Richardson, Ria R. Ghai, Brian Pope, Jamie Yeadon, Betsy Culp, Casey Barton Behravesh, Lixia Liu, Jennifer A. Brown, Leslie V. Boyer

## Abstract

Animal to human transmission of SARS-CoV-2 has not previously been reported in a zoo setting. A vaccinated African lion with physical limitations requiring hand feeding tested positive for SARS-CoV-2 after development of respiratory signs. Zoo employees were screened, monitored prospectively for development of symptoms, then re-screened as indicated, with confirmation by RT-PCR and whole-genome virus sequencing when possible. Trace-back investigation narrowed the source of infection to one of five people. Three exposed employees subsequently developed symptoms, two with viral genomes identical to the lion’s. Forward contact tracing investigation confirmed probable lion-to-human transmission.

Close contact with large cats is a risk factor for bidirectional zoonotic SARS-CoV-2 transmission that should be considered when occupational health and biosecurity practices at zoos are designed and implemented. SARS-CoV-2 rapid testing and detection methods in big cats and other susceptible animals should be developed and validated to facilitate timely implementation of One Health investigations.

---

Although SARS-CoV-2 is primarily transmitted from person-to-person^1^, it is considered zoonotic based on natural infections in a range of mammalian species^2,3^. The broad host range of SARS-CoV-2 is evident from natural infections occurring in zoos, sanctuaries and aquaria, most frequently in big cats^3^, but also in mustelids^4^, non-human primates^5^ and others. Zoo outbreaks typically begin after close contact with an infected zookeeper^6,7,8,9,10^.

One Health investigations have shown transmission of SARS-CoV-2 from people to animals^4,8,9,11,12,13,14^, but examples of animal-to-human transmission are rare^15,16,17,18,19^ and have not been documented in zoo settings.

We report a multispecies cluster of SARS-CoV-2 infections associated with an infected African lion *(Panthera leo)* at a seasonal, mid-sized, Association of Zoos and Aquariums (AZA)-accredited Indiana zoo. The lion’s likely source of exposure was a fully vaccinated, asymptomatic employee, and there was probable forward transmission from the lion to other fully vaccinated employees. To our knowledge, this is the first documented instance of transmission from a zoo animal to a person.

## Methods

### SETTING

The sentinel case occurred in December 2021, while the zoo was closed for the season. The lion was housed alone in a building with an indoor/outdoor enclosure located >30 feet from other animal enclosures. Feeding and veterinary procedures were conducted indoors by dedicated staff with assigned key access. Susceptible species elsewhere in the zoo included a snow leopard *(Panthera uncia)*, Amur tigers (*Panthera tigris altaica)*, Amur leopards (*Panthera pardus orientalis)*, North American river otters (*Lontra canadensis*) and several species of non-human primates. All susceptible animals, including the lion, had received two doses of Zoetis Experimental Mink Coronavirus Vaccine (ZOEMC) between September and November 2021. Animal health was overseen by a full-time on-site veterinarian.

Zoo practice prior to the outbreak included SARS-CoV-2 prevention measures from the Zoo and Aquarium All Hazards Partnership^20,21^. These included suspension of behind-the-scenes tours and close-distance behavioral training for susceptible species. Employees were required to complete the COVID-19 vaccination series, practice social distancing, monitor for COVID-19 symptoms, and when ill to exclude themselves from work. Disinfectant foot baths were placed at enclosure entrances and high-pressure cleaning methods were replaced by methods less likely to aerosolize infectious material. Employees were required to wear surgical masks at all times and nitrile gloves during feedings.

### SENTINEL CASE

The lion was a geriatric (20-year-old) male with chronic renal insufficiency and severe degenerative intervertebral disc disease prohibiting normal range of motion for self-grooming of the caudal body. He was vaccinated with ZOEMC on September 14 and October 7. Routine care included hand feeding twice daily, therapeutic laser treatments of the spine and hindquarters, corticosteroids, gabapentin, renal supplements, and mesenchymal stem cell treatments. On December 18, employees observed coughing, dyspnea, shivering, lethargy, sneezing, nasal discharge, anorexia, and exposed nictitans. Marbofloxacin treatment was initiated. Nasal swabs were collected on December 18 and 23. He was euthanized on December 23 due to declining mobility associated with intervertebral disc disease. The acute clinical signs attributed to SARS-CoV-2 had largely resolved and were not a factor in the euthanasia decision.

### INVESTIGATION

An investigation was initiated on December 18 to identify the source of the lion’s exposure and the potential for forward transmission. (Figure 1)

**Figure 1.**
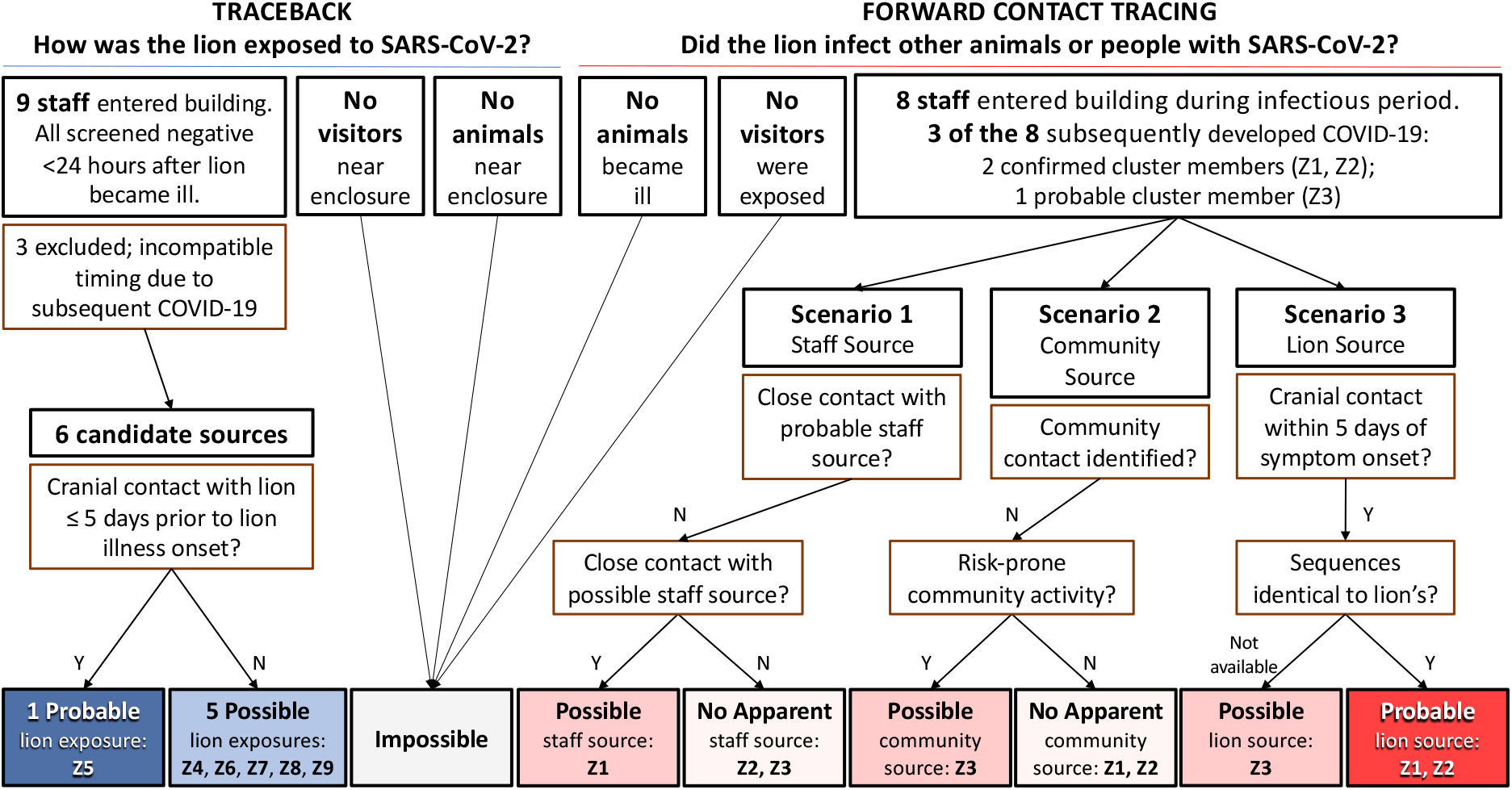
Traceback and Forward Contact Tracing Investigations. The traceback investigation narrowed the potential source of the lion’s SARS-CoV-2 infection to six zoo staff members that had lion contact within 10 days, one of whom (Z5) had cranial contact within 5 days of the lion’s illness onset but did not have close contact with Z1, Z2 or Z3. Possible human sources were identified for Z1 (close occupational contact with Z7) and for Z3 (community activity), although in neither case were these potential sources shown to carry the virus. Cases Z1, Z2 and Z3 all developed symptoms and had confirmed SARS-CoV-2 three days following most recent cranial contact with the sick lion.

A *confirmed cluster-associated case* was defined as a human or animal at the zoo with laboratory evidence for the same strain as the sentinel case by genomic sequencing in a clinical sample. A *probable cluster-associated case* was defined as a human or animal at the zoo with laboratory evidence for SARS-CoV-2 by RT-PCR or rapid antigen test in a clinical sample, but with no genomic sequencing information available.

For both traceback and forward contact tracing investigations, the *exposure period* was assumed to begin 10 days prior to onset of illness, with *probable* acquisition within 5 days prior. The *infectious period* was assumed to begin 2 days before illness and extend 10 days after onset of illness. Any person who entered the enclosure during the lion’s exposure or infectious period was considered to be a *lion contact*. Care provided by lion contacts was classified as *cranial* (feeding, nasal swabbing, 2 feet between human and animal heads) or *caudal* (arthritis care, injections, approximately 9 feet from the lion’s head). *Close contact between people* was defined as proximity of <6 feet for >15 minutes in one day.

### Sentinel case investigation

Nasal swabs from the lion were screened on-site on December 18 by a lateral flow immunoassay validated for human use (Celltrion DiaTrust™ COVID-19 Ag Home Test). Aliquots of samples were sent to the United States Department of Agriculture’s National Veterinary Services Laboratories for confirmatory testing. Necropsy was performed on December 23 at the Michigan State University Veterinary Diagnostic Laboratory.

### Trace back investigation

Nasopharyngeal swabs from all employees exposed to the lion within the 10 days prior to his onset of illness were screened on-site by lateral flow immunoassay on December 18 and 19. Personnel schedules, keeper and veterinary daily reports, maintenance logs, lion treatment schedules, veterinary exam notes, security logs, and social media pages were reviewed for dates and nature of interactions among lion contacts.

### Forward contact tracing investigation

Lion contacts with subsequent symptoms suggestive of infection were screened on-site for SARS-COV-2 by lateral flow immunoassay, within 24 hours of symptom onset. Results were confirmed by RT-PCR (ThermoFisher TaqPath COVID-19 Combo kit) at the Indiana Department of Health (IDOH).

Interviews of infected personnel were performed by an IDOH epidemiologist. These covered symptoms, vaccination history, and exposures to other zoo employees and the public during the 10 days prior to their onset of illness. Location history for activity outside the zoo was verified for these individuals via credit/debit card transaction data where possible. Contact data were cross-referenced with zoo records. Data summaries were deidentified for analysis by authors LB, JAB and RRG.

Relatedness of genomic sequences was analyzed by the authors (LL, BP, JL) by comparison with ten closely related samples collected between August 2021 and February 2022 throughout the state of Indiana. All samples were sequenced in one run utilizing Clear Dx whole-genome sequencing (WGS) SARS-CoV-2 assay with Midnight primers.

The PANGOLIN COVID-19 Lineage assigner (https://pangolin.cog-uk.io/) was used to identify lineage from FASTA files generated by Clear Dx. The phylogenetic tree was constructed with CLC Genomics Workbench version 21.0.3 using the Classical Sequencing Analysis Tools.

### Ethics and Competing Interests

This activity was reviewed by CDC and was conducted consistent with applicable federal law and CDC policy. (See e.g., 45 C.F.R. part 46.102(l)(2), 21 C.F.R. part 56; 42 U.S.C. §241(d); 5 U.S.C. §552a; 44 U.S.C. §3501 et seq.)

All authors have completed the ICMJE uniform disclosure form at www.icmje.org/coi_disclosure.pdf and declare: no support from any organization for the submitted work; no financial relationships with any organizations that might have an interest in the submitted work in the previous three years; no other relationships or activities that could appear to have influenced the submitted work.

### Infection Control

Upon detection of SARS-CoV-2 in the lion, employees providing care for susceptible species at the zoo were required to wear N-95 respirators. Those entering the lion building also used face shields, disposable gowns, shoe covers, and gloves. Lion building access was restricted further and laser treatments were discontinued. Work assignments were altered so that employees providing care to the lion would not come in contact with other susceptible animals.^22^

## Results

### Sentinel case investigation

Evidence of SARS-CoV-2 in the lion was detected by on-site screening on December 18.

Further evidence of SARS-CoV-2 was detected by RT-PCR in nasal swabs from December 18 and December 23 and in nasal turbinate, lung and intestinal tissues obtained at necropsy on December 23. Extracted RNA from both nasal swabs produced high-quality 29,707-nucleotide sequences for genomic sequencing (SARS-CoV-2 Delta variant, AY.103 lineage). Necropsy confirmed intervertebral disc degeneration, chronic renal disease, chronic lower airway disease and severe rhinitis.

### Trace back investigation (Figure 1)

Nine people (employees Z1–Z9) entered the lion building at some time during the lion’s potential exposure period between December 8 and 18. All nine reported being asymptomatic on December 18. Eight had screening tests on December 18 and one had a screening test on December 19; all were negative. Employees Z1, Z2, and Z3 developed COVID between December 21 and 24, with infectious periods that began on or after December 19 (Figure 2).Employee Z5 had cranial contact during the lion’s 5-day “probable” acquisition period. Employees Z4, Z6, Z7, Z8 and Z9 had lion contact 6-10 days before onset of the lion’s illness and/or caudal activities exclusively.

**Figure 2.**
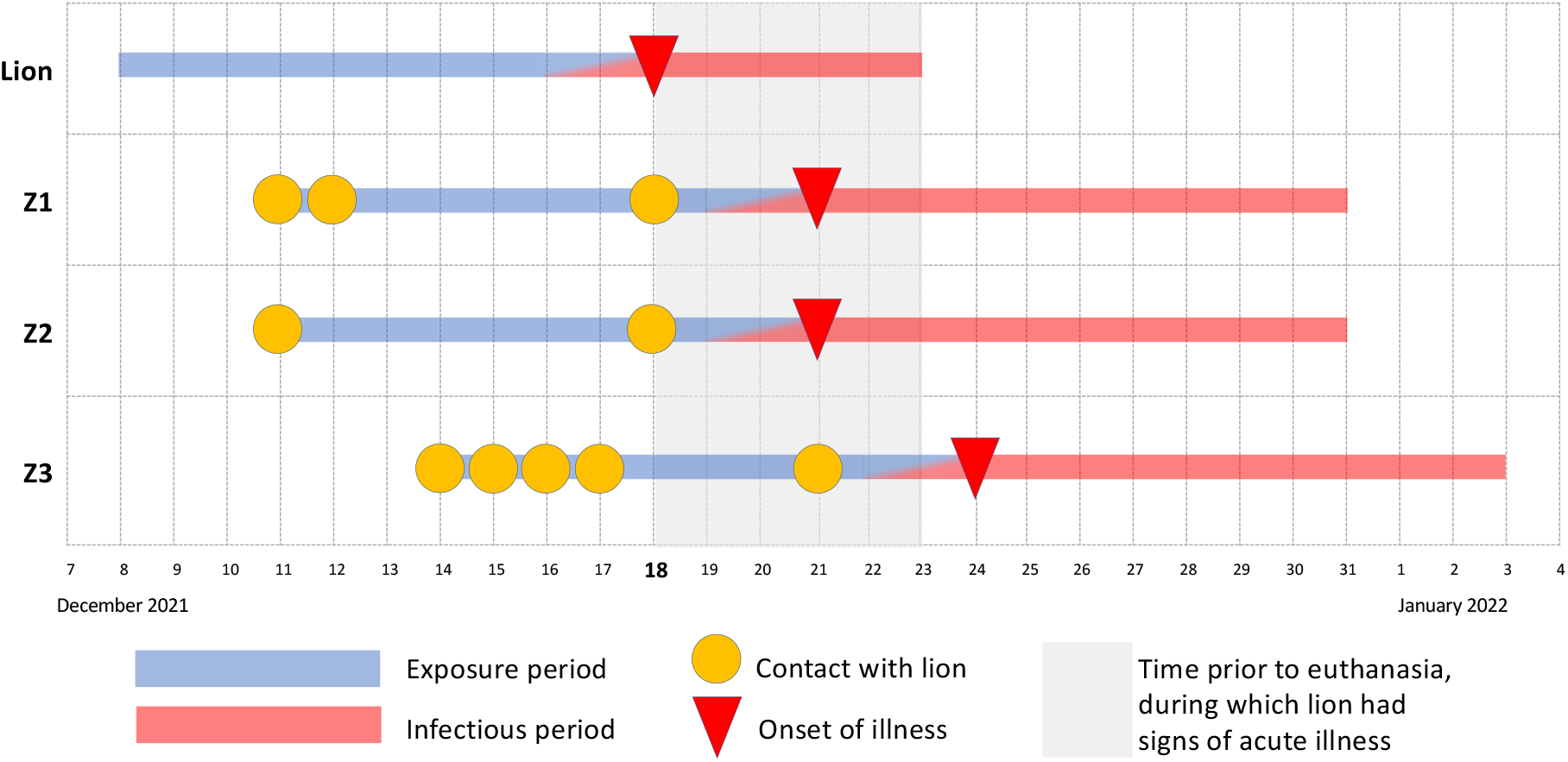
Timeline. The lion’s likely exposure period was December 8–17 and his infectious period was December 16 through euthanasia on December 23. During the lion’s infectious period, Z1 and Z2 each had a single day of cranial contact with him, coinciding with the day of the lion’s onset of illness. Z3 had cranial contact with the lion on three occasions during his infectious period. Note the lack of overlap between Z1 and Z2’s infectious period and the lion’s exposure period, and between Z3’s infectious period and Z1 and Z2’s exposure periods.

### Forward contact tracing investigation (Figure 1)

No additional animals at the zoo developed illness suggestive of SARS-CoV-2.Eight people entered the lion building during the lion’s infectious period, December 16 through 23. Z1, Z2 and Z3 were fully vaccinated and previously healthy employees, each of whom was involved in cranial care 3 days before developing upper respiratory symptoms. Each of these three was positive on rapid antigen test within 24 hours of symptom onset, with subsequent RT-PCR confirmation of SARS-CoV-2. Samples from Patients Z1 and Z2 were suitable for sequencing; the sample from Z3 was of insufficient quality for sequencing.

The sequences generated from lion samples on December 18 and 23 were identical to those from Z1 and Z2 samples. The 10 community comparators varied from the cluster sequences by 12 to 24 nucleotides. Genetic distances among sequences are indicated in Figure 3.

**Figure 3.**
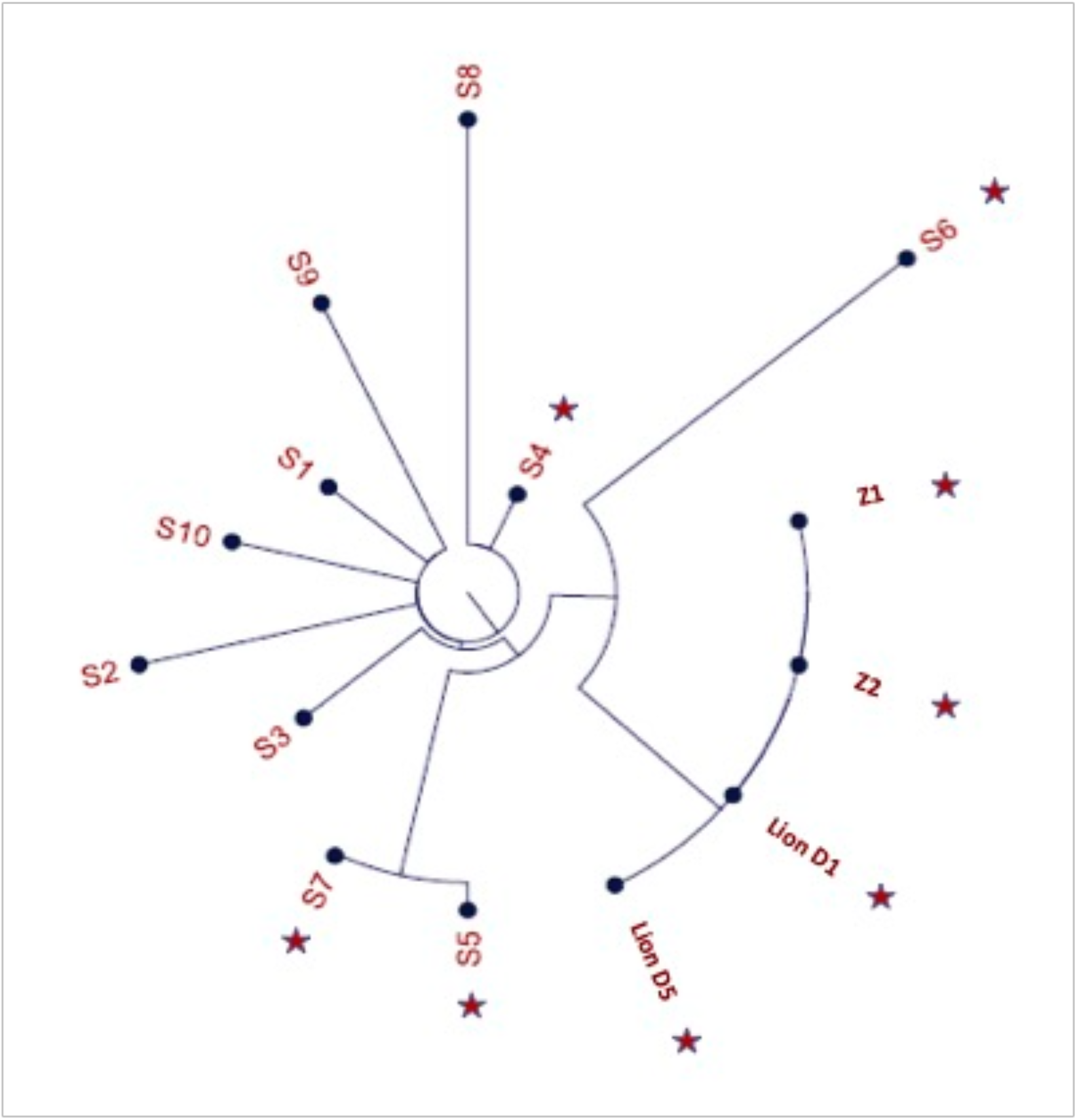
Phylogenetic Tree. SARS-CoV-2 Delta variant, AY.103 lineage genome sequences from the lion (day 1 and day 5) and zoo employees (Z1 and Z2) are shown in comparison with reference sequences from COVID-19 patients from 7 counties in Indiana, August 2021-February 2022. Reference sequences are labeled chronologically as S1 to S10. Stars denote specimens collected in December 2021.

Employees Z5, Z8 and Z9 had no close contact with Z1, Z2 or Z3 during their exposure periods. Z4 had close contact with Z2 on December 18, the day that both had negative screening tests. Z6 entered the lion enclosure at the same time as Z3, but maintaining distance >6 feet, 9 days before Z3’s symptom onset. Z7 had close contact with Z1 on December 11, 12 and 18. Z7 worked in the lion enclosure with Z2, but this contact took place a full 10 days prior to Z2’s onset of symptoms and was at >6 feet for <15 minutes. Z7 also worked in the lion enclosure with Z3, for <15 minutes and at a distance of >6 feet 8 days prior to Z3’s onset of symptoms.

Patient Z3 had no known exposures outside of the zoo but did participate in community social activities with risk for unrecognized transmission of SARS-CoV-2 on several occasions during the 10 days prior to onset of symptoms.

## Discussion

This multispecies cluster of SARS-CoV-2 included three confirmed cases (two human, one felid) and one probable case (human). The identical genomic sequences detected in samples from the confirmed cases demonstrate that the infections were acquired in a common setting. Given that the zoo was closed to visitors, the source of infection for the sentinel case was almost certainly one of 6 asymptomatic employees who tested negative on the day of the lion’s diagnosis and who subsequently reported no signs of illness. Among these 6, Employee Z5, the only person who had cranial contact within 5 days before the lion’s onset of illness and who did not later develop symptomatic COVID-19, was the most likely source of the lion’s infection.

To determine whether lion-to-human transmission took place, we considered 3 scenarios (Figure 1), below.

*Scenario 1: Zoo employees acquired infection from the same human source as the lion*. Although employee-to-employee transmission could not be ruled out on an individual basis, no pathway of transmission could be identified that explained all probable cases within the cluster.

*Scenario 2: Zoo employees acquired infection from an unrelated community source*. Employees Z1 and Z2 had no other identified sources of SARS-CoV-2 transmission from the community and samples from both yielded sequences genetically identical to the lion. For Z3, whose viral genome was unknown, community acquisition is possible.

*Scenario 3: At least one zoo employee acquired infection from the lion*. During cranial-end procedures, the lion breathed, roared and coughed within arm’s length of staff. Each person that developed symptoms after the lion was involved in cranial-end care while the lion was sick, and each developed symptoms 3 days following that exposure. Lion-to-human transmission of SARS-CoV-2 is therefore the most probable explanation for their infections.

Our investigation strongly suggests that lion-to-human transmission took place in at least one, and up to three, instances. This is important for at least two reasons. First, animal-to-person transmission of SARS-CoV-2 is an occupational health risk for veterinary and animal care staff that interact closely with susceptible animals. Second, transmission occurred despite an up-to-date SARS-CoV-2 vaccination history in every individual involved, including the person(s) that likely transmitted the virus to the lion, the lion, and the person(s) that likely became infected from the lion. While SARS-CoV-2 transmission from vaccinated people to vaccinated zoo animals has been documented previously^23^, our results show that transmission can also occur from vaccinated zoo animals to vaccinated people. While many human vaccine efficacy studies indicate reductions in disease severity of COVID-19 following vaccination^24,25^, further research is needed to determine if the same is true of zoonotic SARS-CoV-2 transmission among vaccinated people and animals.

The unique setting minimized the number of potential exposure pathways required to reach this conclusion. Initiation of the investigation and heightened safety measures on the day of sentinel case discovery facilitated analysis and contained spread. Timely screening, prospective monitoring for symptom development, cross-referencing of occupational and interview data, and genomic sequencing combined to allow inferences of causality.

Swabbing leonine nasal passages for SARS-CoV-2 diagnosis has been reported^26^, but neither the sampling method nor the use of tests designed for human diagnosis has been formally validated. Rapid testing methods applicable to animals should be developed and validated to allow for timely implementation of biosecurity measures.

This report is subject to at least three limitations: first, only a limited number of community samples was available for the sequencing analysis. Second, rapid tests may not reliably establish SARS-CoV-2 negative status. Third, the distance associated with close contact and exposure and infectious periods for lions were assumed to be similar to those for humans, although these have not been established.

Timely human-animal-environmental assessments and implementation of appropriate biosafety interventions are essential for both animal and human health during community outbreaks of SARS-CoV-2. A One Health approach, which connects the health of people, animals, and environment^27^, should be used to respond to susceptible zoo animals, particularly those requiring close human contact, that develop clinical signs compatible with SARS-CoV-2 infection. Close contact with large cats should be considered a risk factor for bidirectional zoonotic SARS-CoV-2 transmission, regardless of prior immunization. This may be especially true in geriatric animals or those with underlying health conditions.

Audrey Siegrist, DVM, is a practicing zoo animal veterinarian currently affiliated with the Potawatomi Zoo. Her research interests include envenomation of captive animals by native reptiles and arthropods and One Health investigation of infectious disease outbreaks.

## Data Availability

All data produced in the present study are available upon reasonable request to the authors

